# Development and Validation of Minimal Disease Activity, Disease Flare, and Minimal Clinically Important Difference for Children with Chronic Nonbacterial Osteomyelitis Using a Consensus and Data-Driven Approach

**DOI:** 10.64898/2025.12.04.25340995

**Authors:** Farzana Nuruzzaman, Natalie Rosenwaser, Xing Wang, Jonathan D. Akikusa, Matthew L. Basiaga, Ava Klein, Ian Muse, Erin Balay-Dustrude, Megan Nguyen, Emily Deng, Aleksander Lenert, Lindsey Bergstrom, Fatma Dedeoglu, Bin Huang, Jenna King, Sivia Lapidus, Tzielan C. Lee, Cassandra M. Levesque, Lillian Lim, Kimberly Martin, Elizabeth Murray, Melissa S. Oliver, Karen B. Onel, Seza Ozen, Lauren Potts, Sara M. Stern, Robin Villaverda, Evelyn Y. Wu, Ronald M. Laxer, Polly J. Ferguson, Daniel J. Lovell, Yongdong Zhao, the CHOIR network and CARRA CNO workgroup

## Abstract

**Objectives:** To create definitions for minimal disease activity (MDA), flare, and minimal clinically important difference (MCID) for chronic nonbacterial osteomyelitis (CNO). It is necessary to establish these criteria prior to starting clinical effectiveness trials for therapies in CNO.

**Methods:** Three separate cohorts samples from an international observational CNO registry were presented to a panel of twenty experts in CNO, including 7 patients/caregivers via online and in-person voting using nominal group technique to define MDA, flare, and MCID, respectively. Experts classified the cases in each cohort as meeting the state for that specific cohort, ie. MDA or not, or flare or not, or meeting MCID or not. A consensus of ⩾80% was required.

**Results:** General surveys identified the most important variables to include to define MDA, flare, and MCID. Clinical improvement of 30% or more in these critical parameters and improvement of CNO Clinical Disease Activity Score (CDAS) by at least 3 were considered meaningful by providers and consensus data.

Conclusions

This study provides the preliminary definitions of MDA, disease flare, and MCID in CNO which can serve as targets and potential outcomes from treatments. These definitions must now be validated in other cohorts and tested in clinical trials.

**Key Messages:** *WHAT IS ALREADY KNOWN ON THIS TOPIC:*

- Measurements of minimal disease activity (MDA), worsening disease activity (‘flare’), and minimally clinically important differences (MCID) in CNO are not well-established. For successful execution of clinical trials to identify effective treatment strategies in patients with CNO, standardized definitions of these terms are necessary.

*WHAT THIS STUDY ADDS:*

- Using data from an international real-world registry of CNO patients, we developed and validated quantitative definitions for MDA, flare, and MCID.

*HOW THIS STUDY MIGHT AFFECT RESEARCH, PRACTICE OR POLICY:*

- Definitions of MDA, flare, and MCID will serve as potential outcomes in future clinical trials to study effectiveness of therapies in CNO.

## Introduction

Chronic nonbacterial osteomyelitis (CNO) is an autoinflammatory bone disease of unknown cause that can result in bone destruction/deformity, persistent bone pain and pathological fractures. While recent classification criteria have been developed, the diagnosis of CNO is often one of exclusion (1). Consensus treatment plans have been developed for children with CNO who have an NSAID-refractory course and/or with active spinal lesions, to limit treatment practice variation and enable comparative effectiveness trials (2). However, due to lack of randomized control trials (RCTs) from international and multi-ethnic cohorts, treatment is often empirical and based on clinical experience, expert opinion, or retrospective observational data (3–9). This impacts patients negatively, particularly those with refractory disease or other associated conditions. A recent international conference involving clinicians, researchers, patients, and public stakeholders identified that medication trials are among the top research priorities in CNO (10). In order to conduct meaningful clinical research, disease states such as minimal disease activity (MDA), disease flare, and minimal clinically important differences (MCID) must be well defined and validated (11–13). It is critical to involve patients/caregivers to define these states as these terms are linked to patient-reported outcomes, adaptation to diagnosis, and treatment decisions and response (14, 15). Remission or inactive disease over a period of time is the ultimate goal to achieve for all patients with rheumatological diseases. However, MDA is often used as a part of the treat to target approach because it represents a level of low disease that is acceptable for both patients and providers that may be more readily achievable at a single time point. Additionally, it is equally important to define the point of loss of efficacy of treatment at which the patient experiences a disease flare during RCTs, longitudinal observational studies and post-marketing studies. A third critical outcome measure to define is MCID, an estimation of the smallest change in measurements that signifies an important difference/improvement of a disease caused by an effective medication. We are building the framework on which to design future clinical trials in CNO to evaluate and compare medication efficacy which can further guide treatment decisions for patients with these pillars in place. Objectives of this study were to develop and validate definitions for MDA, disease flare, and MCID in CNO.

## Methods

### General Survey

A general survey regarding priorities for defining MDA and flare was sent to CNO patients/caregivers through a Facebook CNO support group (∼6500 members) and to providers via the Childhood Arthritis and Rheumatology Research Alliance (CARRA) international CNO Workgroup (∼180 members) between December 20, 2024 and February 26, 2025 **(Supplement 1).** Additional questions regarding MCID were also sent to providers only **(Supplement 2)**. The survey was voluntary and anonymous. Informed consent was obtained online immediately prior to initiating the survey.

### Patient and caregiver involvement

Patients and caregivers who are active participants in the CARRA CNO Workgroup meetings were engaged as research partners and were involved in the design and conduct of this research. They helped determine feasibility, prioritized research questions, survey development, and helped recruit patients to complete the survey. Invited patients and caregivers also participated as members of the expert voting panels.

### Expert Panel Participants

An expert panel was convened, including pediatric rheumatologists from the CARRA CNO Workgroup, patients with CNO, and caregivers of pediatric patients with CNO from Facebook. A purposive sample of providers, patients and caregivers was identified, and individuals were contacted and invited to participate by email. The sampling aimed to recruit people who would be familiar with CNO either through clinical management of patients with CNO, researching in CNO, or living with CNO and caring for patients with CNO. Consent was waived per Seattle Children’s institutional review board.

### Chronic nonbacterial osteomyelitis international registry (CHOIR)

The <u>ch</u>ronic nonbacterial <u>o</u>steomyelitis international <u>r</u>egistry (CHOIR) is a multisite international observational longitudinal cohort study of children with CNO that was established by the CARRA CNO workgroup in 2018 (<u>ClinicalTrials.gov</u> Identifier: <u>NCT04725422</u>) (16). The CHOIR registry contains data from >3000 real-world patient visits that can be leveraged for a data-driven approach.

### Chronic nonbacterial osteomyelitis clinical disease activity score (CDAS)

The CNO clinical disease activity score is the sum score of patient pain (scored on a 0-10 Likert scale), patient/parent global assessment (PAGA) (scores on a 0-10 Likert scale) and clinically active CNO lesion count defined as focal tenderness, and/or swelling, and/or warmth in addition to patient’s report of pain at a known CNO lesion site. The CNO CDAS was developed and validated for disease monitoring and assessment of treatment effectiveness (16).

### Cohort Creation used for Definition of MDA and Flare

Based on survey responses, key parameters from CHOIR were extracted to create two cohorts of 600 cases each to define MDA and flare (**Table 1**). Patient pain level (0–10), PAGA (0–10), physician global assessment (PHGA) (0–10), clinical lesion count, type of MRI, total MRI lesion count, active spinal lesion on MRI, active sacroiliitis on MRI, erythrocyte sedimentation rate (ESR), and C-reactive protein (CRP) were extracted. Missing variables were imputed.

**Table 1.** Characteristics of Cohorts Used to Develop Definitions of Minimal Disease Activityand Flare in CNO

|  | MDA Cohort (n=600) | Flare Cohort (n=573) |
| --- | --- | --- |
| Age: in years, median [IQR] | 12.7 [10.2, 15.1] | 12.8 [10.3, 15.1] |
| Sex (n, %) | Female: 320 (55)<br>Male: 261 (45) | Female: 309 (56)<br>Male: 248 (45) |
| Extracted Parameters |  |  |
| Pain Level (0-10): median [IQR] | 1 [0, 4] | 1 [0, 4] |
| Patient global assessment (PAGA) (0-10): median [IQR] | 2 [0, 5] | 2 [0, 5] |
| Physician global assessment (PHGA) (0-10): median [IQR] | 1 [0, 2] | 1 [0, 2] |
| CNO clinical lesion count: median [IQR] | 0 [0, 1] | 0 [0, 1] |
| Active clinical arthritis: n (%) | No: 505 (84)<br>Yes: 53 (9) | No: 478 (83)<br>Yes: 53 (9) |
| MRI type: n (%) | Whole Body MRI: 531 (89)<br>Regional with Spine: 12 (2)<br>Regional without Spine: 57 (10) | Whole Body MRI: 504 (88)<br>Regional with Spine: 12 (2)<br>Regional without Spine: 57 (10) |
| Total MRI lesion count: median [IQR] | 3 [2, 4] | 3 [2, 4] |
| Active spinal lesion on MRI: n (%) | No: 532 (89)<br>Yes: 68 (11) | No: 505 (88)<br>Yes: 68 (12) |
| Active sacroiliitis on MRI: n (%) | No: 571 (95)<br>Yes: 29 (5) | No: 544 (95)<br>Yes: 29 (5) |
| Erythrocyte Sedimentation Rate (ESR): n (%) | Abnormal: 105 (18)<br>Normal: 495 (83) | Abnormal: 104 (18)<br>Normal: 469 (82) |
| C-Reactive Protein (CRP): n (%) | Abnormal: 68 (11)<br>Normal: 532 (89) | Abnormal: 67 (12)<br>Normal: 506 (88) |
| CNO Disease Activity Score (CDAS)*: median [IQR] | 4 [1, 10] | 5 [1,10] |

\*CDAS: Sum of patient pain score (0-10), PAGA (0-10), and total number of clinically active CNO lesions defined as focal tenderness, and/or swelling, and/or warmth

An expert panel of 13 international pediatric rheumatologists and 7 patient/caregiver research partners reviewed the cases individually. Each case was reviewed by 5 panelists. 10-20 common cases were reviewed by all 20 panelists. Cases which did not reach consensus agreement were reviewed by ≥5 reviewers at an in-person consensus meeting held in Denver, Colorado on April 2-3, 2025. For each cohort, reviewers were asked to vote whether a clinical scenario was considered MDA or flare, respectively. For the purpose of this exercise, panelists were provided the following information:

● “Minimal disease activity (MDA) refers to a state when a patient feels completely or reasonably well where the patient/caregiver and medical providers do not feel the need to change medical therapies at that time.”
● “Flare refers to a state when a patient feels so unwell that a patient and medical provider need to restart medications (after the medication has been stopped during medicated remission) or escalate current medical therapy if already on treatment.”

Consensus was defined as ≥80% of agreement among experts using nominal group technique. A multivariate general linear model (GLM) was performed using 80% of data to develop the model and cross-validation was used to verify the results. Decision tree analysis was performed.

### Cohort Creation used for Defining MCID

Similar to cohorts used to define MDA and flare, based on Delphi survey results, 664 cases were identified from CHOIR to create a cohort of paired visits with key parameters to define MCID **(Table 2**). Patient pain (0–10), PHGA (0–10), clinical lesion count, total MRI lesion count, active spinal lesion on MRI, active sacroiliitis on MRI, ESR, CRP, new spinal lesion, new non-spinal lesion were extracted from paired visits. Inclusion criteria from the baseline visit were: MRI lesion count and the sum of patient pain and PHGA as well as the sum of PAGA and PHGA are greater than 0. The interval between two visits was between 3 and 12 months. At least one or more values at the follow up visits have improved while no more than 1 value worsened.

**Table 2:** Characteristics of Cohorts Used to Develop Definitions of Minimal Clinically Important Difference in CNO

| n= 664 | MCID Cohort (baseline visit)* | MCID Cohort (followup visit) | p-values^ |
| --- | --- | --- | --- |
| Age: median [IQR] | 12 [9.4,14] |  |  |
| Sex: n (%) | Female: 391 (60)<br>Male: 257 (40) |  |  |
| Extracted Parameters |  |  |  |
| Pain Level (0-10): median [IQR] | 5 [3, 6] | 2 [0, 4] | <0.0001 |
| Patient global assessment (PAGA) (0-10): median [IQR] | 5 [3, 6] | 2 [0, 4] | <0.0001 |
| Physician global assessment (PHGA) (0-10): median [IQR] | 2 [1, 4] | 1 [0, 2] | <0.0001 |
| CNO clinical lesion count: median [IQR] | 1 [0, 2] | 0 [0, 1] | <0.0001 |
| Total MRI lesion count: median [IQR] | 3 [2, 5] | 2 [1, 3] | <0.0001 |
| Active spinal lesion on MRI: n (%) | No: 510 (77)<br>Yes: 154 (23) | No: 588 (89)<br>Yes: 76 (11) | <0.0001 |
| Active sacroiliitis on MRI: n (%) | No: 544 (82)<br>Yes: 120 (18) | No: 610 (92)<br>Yes: 54 (8) | <0.0001 |
| New spinal lesion on MRI: n (%) | N/A | No: 647 (97)<br>Yes: 17 (3) | N/A |
| New non-spinal lesion on MRI: n (%) | N/A | No: 366 (55)<br>Yes: 298 (45) | N/A |
| Erythrocyte Sedimentation Rate (ESR): n (%) | Abnormal: 205 (31)<br>Normal: 459 (69) | Abnormal: 73 (11)<br>Normal: 591 (89) | <0.0001 |
| C-Reactive Protein (CRP): n (%) | Abnormal: 75 (11)<br>Normal: 589 (89) | Abnormal: 30 (5)<br>Normal: 634 (96) | <0.0001 |
| CNO Disease Activity Score (CDAS)#: median [IQR] | 10 [7, 14] | 4 [1, 8] | <0.0001 |

\*Inclusion criteria for baseline visit were: MRI lesion count and the sum of patient pain and PHGA as well as the sum of PAGA and PHGA are >0
^p-values calculated using Wilcoxon signed rank test for continuous variables and McNemar's test for categorical variables
#CDAS: Sum of patient pain score (0-10), PAGA (0-10), and total number of active CNO clinical lesions defined as focal tenderness, and/or swelling, and/or warmth

Members of the expert panel reviewed the cases to determine if they showed a minimal clinically important difference between the paired visits. For the purposes of this exercise, MCID was considered “a way of measuring if a certain medication is working” or “the smallest change that we consider meaningful in CNO bone and joint disease after starting a treatment” by the voting panel. Cases which did not reach consensus (80% agreement) were evaluated by 5 or greater reviewers at the in-person consensus meeting. Using the data from these cases, a decision tree analysis method was used to determine the cutoff point that best differentiated patients with and without MCID, and the model was validated using 10-fold cross-validation to ensure generalizability.

## Results

### Survey Results

A total of 252 participants (133 caregivers, 39 patients and 80 providers) consented and completed the survey. The survey was completed by respondents from 5 continents (77% from North America). A total of 80 providers (44% response rate) from 5 continents completed the survey. Most providers (90-96%) had >5 years of experience treating children with CNO with ≥5 patients per year. Among patients/caregivers, 91% were diagnosed within 10 years, 98% had taken medications for CNO, and 85% had experienced a flare. The frequency of items deemed important in defining MDA and flare selected by two groups was discrepant on the survey (**Figure 1**). The frequency of items selected as parameters important in determining MCID was listed, and the absolute and relative change were reported (**Figure 2**).

**Figure 1:**
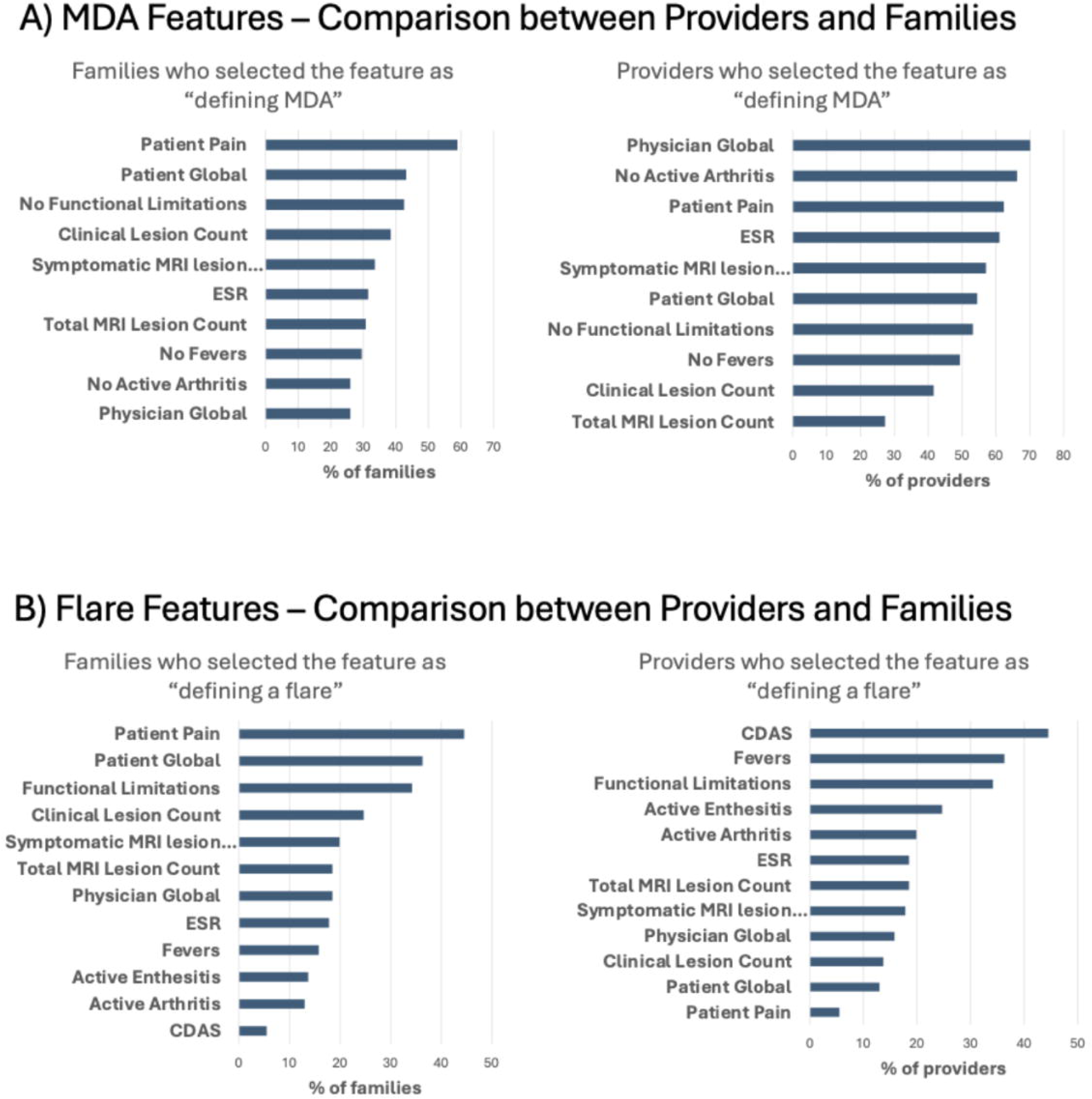
Provider and Patient/Caregiver Family Priorities in Defining Minimal Disease Activity and Flare in CNO Delphi Survey Results

**Figure 2:**
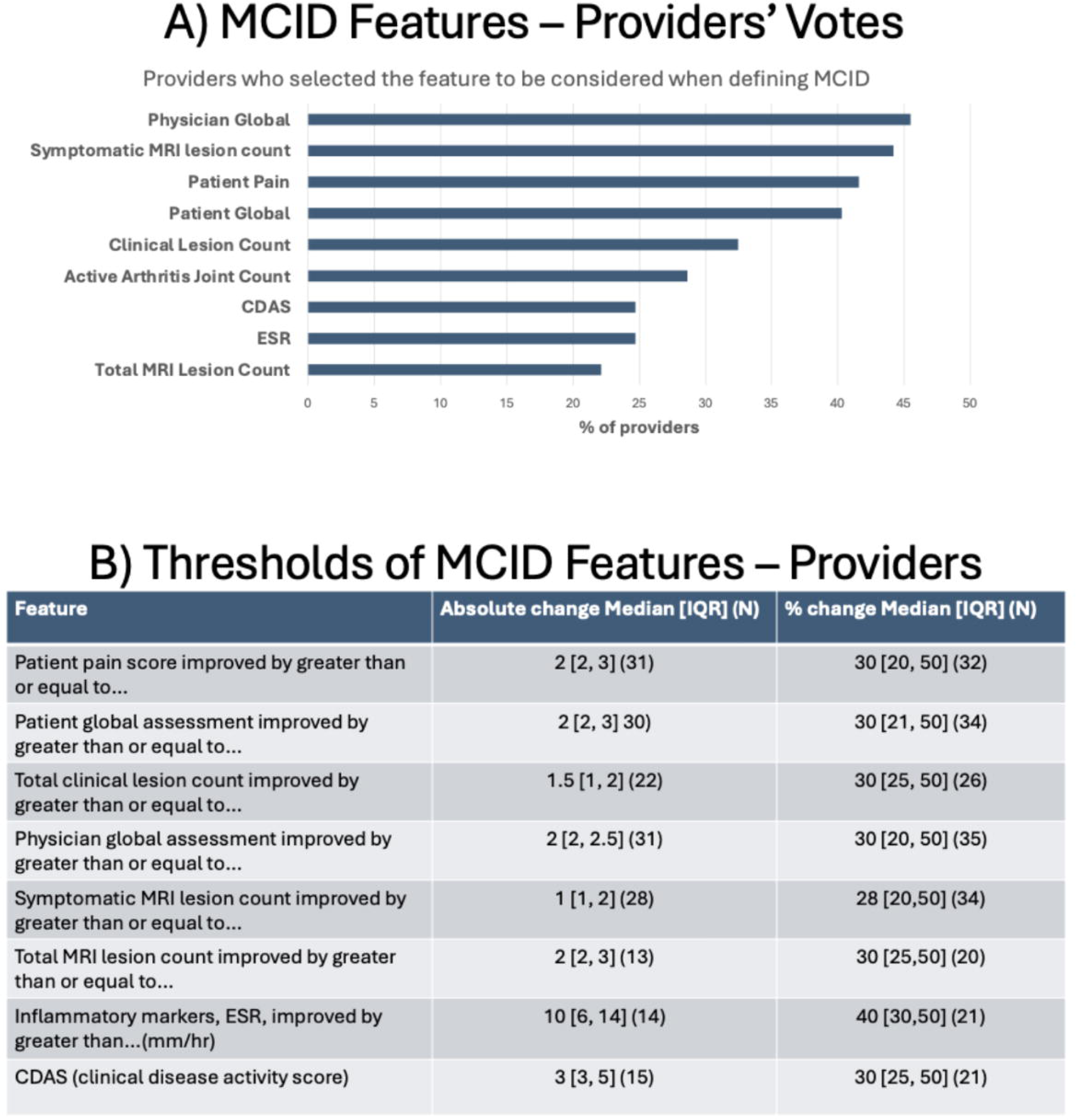
Provider Priorities in Defining Minimal Clinically Important Difference in CNO Delphi Survey Results: Frequencies and Thresholds

### Expert Panel Results

The purposive sampling process included 21 physicians and 19 patients/caregivers. In total, 13 physicians (excluding 4 moderators, DL, FN, NR, YZ) and 7 patients/caregivers agreed to participate in the case definition process. All participants were invited to attend a face-to-face consensus meeting. Twelve physicians and six patients/caregivers attended the consensus meeting in all. Some (n= 4) participants chose to participate via Zoom video teleconferencing. One person from each group (both providers) agreed to participate but withdrew (KO, SO) during voting for MCID, leaving 6 participants in each group for the remainder of patient profiles after case #76. All participants are involved in CNO care and/or research or have lived experience or care for a person with CNO.

### Voting Results for MDA/Flare Cohorts

253 cases were classified as having MDA and 324 were classified as not having MDA whereas 23 cases were inconclusive. GLM analysis showed that pain level (p<0.001), PHGA (p=0.012), PAGA (p=0.005), clinical lesion count (p=0.001) and MRI lesion count (<0.001) were significantly associated with MDA classification. Of the flare cohort, 404 were classified by consensus as flare, 159 as non-flare and 10 were inconclusive. Pain level (p<0.001) and MRI lesion count (p=0.049) were associated with flare designation. Cases with presence of either sacroiliitis or spinal lesions on MRI or active clinical arthritis were consistently voted as flare and as non-MDA (p<0.001). The median [IQR] clinical disease activity score (CDAS) was 0 [0,2] in cases with MDA compared to 10 [6, 12] in those without MDA (p<0.001) with a threshold CDAS ≤3 for being categorized as MDA. The accuracy of the MDA model without CDAS and with CDAS was 0.77 and 0.89, respectively. The median [IQR] CDAS was 8 [4, 12] in cases with flare, compared to 0 [0, 2] in those without flare (p<0.001) with a threshold CDAS ≥3 for being categorized as flare. The accuracy of the flare model without CDAS and with CDAS was 0.78 and 0.87 respectively. Decision pathways were developed (**Figures 3 and 4**).

**Figure 3:**
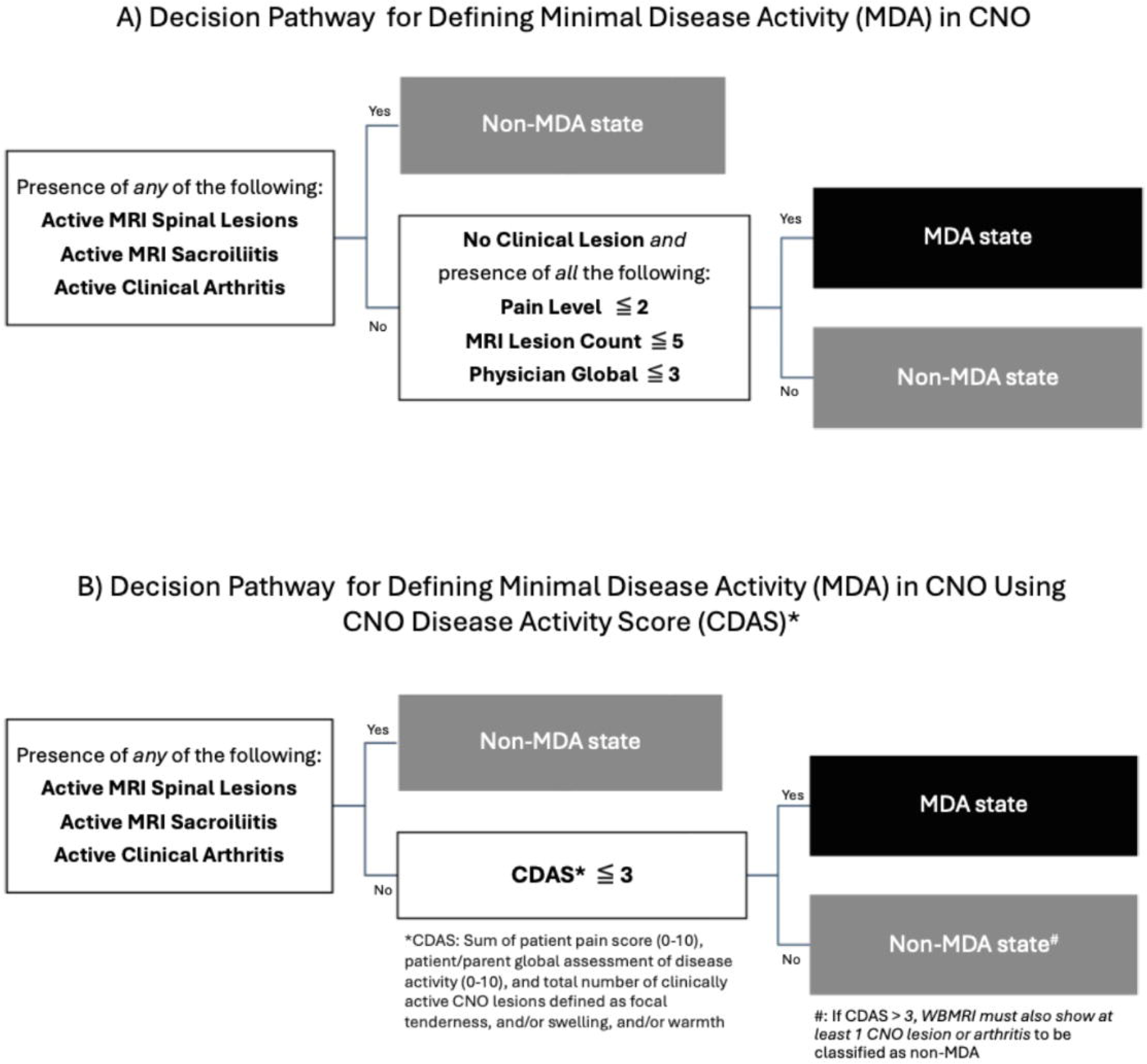
Decision Pathways for Defming Minimal Disease Activity in CNO

**Figure 4:**
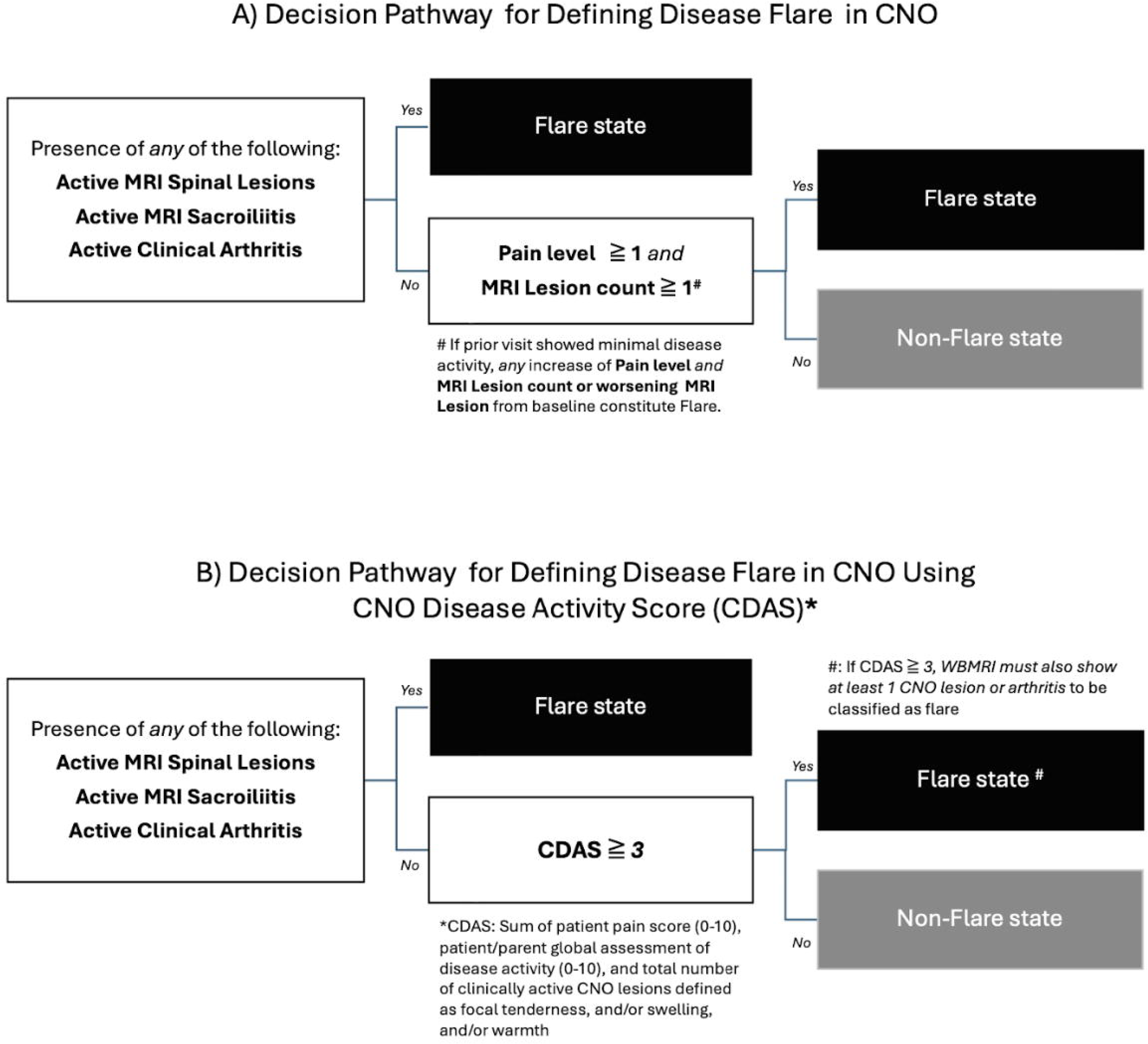
Decision Pathways for Defining Disease Flare in CNO

### Voting Results for MCID

Consensus was reached by 638 (96%) of 664 paired visits, with 423 classified as MCID, 215 as no MCID and 26 inconclusive. A generalized linear model analysis showed that the change of pain level (scale 0-10, p=0.03), change of MRI lesion count (p<0.001) and change of active spinal lesion on MRI (p<0.02) were associated with the classification of MCID. The median [IQR] change of clinical disease activity score (CDAS) was -7 [-11,-4] in cases with MCID, compared to -2 [-5, 0] in those without MCID (p<0.001). The improvement of CDAS by ≥3 was categorized as meeting MCID. Decision trees based on all parameters or CDAS alone with accuracy of the model as of 0.73 and 0.71, respectively are reported in **Figure 5**.

**Figure 5:**
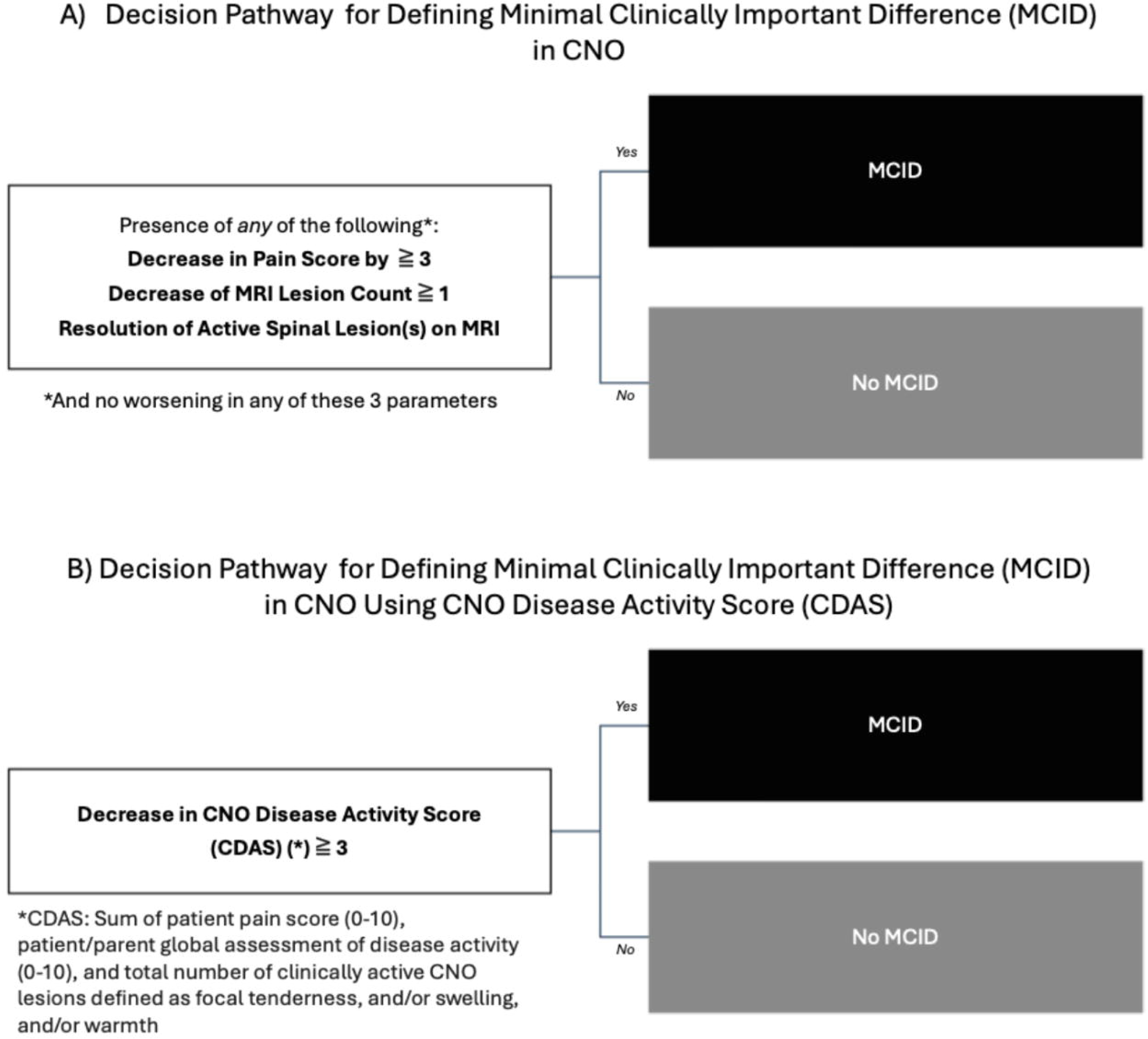
Decision Pathways for Defming Minimal Clinically Important Difference (MCID) inCNO

## Discussion

In this study, we sought to determine criteria that corresponded with MDA, disease flare and MCID in patients with CNO. These definitions are the cornerstones on which to build observational studies and future clinical trials for this condition. Due to potential mimickers, CNO itself is a challenging disease to diagnose. Additionally, it is considered a relatively rare disease, though recognition may be increasing due to availability of a more unified classification criteria recently promoted by EULAR/ACR (2). Additionally, CNO has a wide range of clinical presentations, from a unifocal lesion that responds to first-line therapy with NSAIDs, to a chronic relapsing disease with multiple lesions that requires therapies with second-line agents including TNF inhibitors and/or bisphosphonates. Discrepancies between histological findings, MRI characteristics, and patient-or-parent reported symptoms complicate objective disease activity evaluation. Despite these challenges, we successfully developed pathways for each of these critical terms using clinical criteria and CNO composite disease activity (CDAS) scores in a collaborative, data-driven approach. The CARRA CNO workgroup had introduced the CDAS as a tool to estimate disease activity which focuses on patient and parental considerations and clinical lesions (14). The relative importance of the presence of spine and sacroiliac lesions was evident in the final pathways for all definitions. Consensus was sought on definitions for MDA, flare, and MCID that can be recommended for use in future clinical trials and further validated in other datasets and long-term outcome databases. Further validation is needed before these definitions are ready to be used to guide decisions in individual patient care.

Other necessary components for future successful clinical trials in CNO would be a centralized image scoring tool to ensure that readings of MRIs are consistent and reproducible. A web-based ChRonic nonbacterial Osteomyelitis Magnetic Resonance Imaging Scoring: (CROMRIS) tool has been proposed and preliminarily validated to be used as a framework for this tool (17). A proposed trial to compare adalimumab to bisphosphonates has been described as a needed trial to generate evidence to help with patients obtaining approval for medications and avoid delays in treatment (18). The distribution of characteristics of children with CNO receiving tumor necrosis factor inhibitors and those receiving bisphosphonates in CHOIR was comparable and balanced across groups suggesting that it is feasible to compare the effectiveness of these two medications within CHOIR as a future study (14). Thus, development of the parameters of MDA, flare and MCID is a timely necessity to advance CNO research.

We aimed to define *minimal disease activity* in CNO instead of *inactive disease activity* or total absence of signs/symptoms of disease activity as the former has been suggested as a more practical target for clinical care (11). MDA also implies a patient’s clinical state at a single time point. MDA is also distinct from remission in that remission is a state of inactive disease over a set period of time. CDAS as a composite measure was assessed in the analysis phase as described in the results section because individual component data were available. Although the CDAS score was relevant, because it is a composite measure (of patient pain, PAGA, and clinical CNO lesion count), the voting panel members were reassured that all components of the CDAS were included in the patient profiles rather than voting on specific CDAS scores. Panelists made it clear that presence of sacroiliac joint (SIJ) involvement in itself necessitated treatment. Thus, it would drive them to consider a case as not in MDA regardless if individuals considered the SIJ involvement to be part of the clinical profile of CNO or whether it is thought to be a part of manifestations of coexistent juvenile idiopathic arthritis separate from CNO.

In prior literature, *flare* in CNO has been defined as an increase in disease activity either clinically (increase in pain levels), radiologically (increase in number or size of MRI-confirmed lesions), or some combination of both (19–21). For the purposes of consensus, we used the convention that flare represented enough disease worsening that it is clinically appropriate to restart or escalate CNO treatment. During the nominal group technique discussion panelists were advised that clinical lesions indicated evidence of active inflammation (i.e. tender upon palpation, reversible swelling, or warmth) found on objective clinical exams. Reversibility of swelling was important to differentiate from swelling resulting from bony hypertrophy due to untreated disease that is not amenable to improvement by therapy. During voting, it became evident that pain level became a preeminent parameter to consider during the decision-making process in assessing relapse. Panelists who were physicians became more sensitive to pain levels after hearing perspectives from panelists who were patients/caregivers, and the level of “acceptable” pain shifted during the voting process.

There have been no prior studies on MCID in CNO, thus, several concepts were discussed by the voting panel. MCID was proposed to be used as an outcome in intervention studies to assess minimal improvement to be considered responsive to the treatment (22). It was noted that this degree of difference between the baseline and follow-up appointment needs to rise above the inherent variability in the disease symptoms and signs that occurs without any change in treatment. This point was most resonant with the patients and parents who had observed this variability firsthand. Pain was a very important, if not the most important, parameter being used in the consideration of clinical change as was highlighted in the discussions above regarding disease flare. Eventually there was convergence between the opinions of patients/parents and providers about the change in level of pain deemed significant enough to drive the decision on MCID. However, all participants noted that the convention used in this study, that all pain to be considered in patient profiles was to be due to CNO, was not in keeping with the more complex situation in routine clinical care in which other factors often play a major role in patient reported pain. It was thought that, in the future, emphasis should be placed on asking the parent or patient to report “CNO related pain” rather than a total pain score because although challenging, many patients could do this fairly accurately.

Discussion of specific cases highlighted the importance of new or persistent spinal lesions even in the situation of other aspects of the disease being well-controlled or minimally active. Resolution of sacroiliac or spine lesions alone often was sufficient to lead the panel voting in favor of MCID even when all other components were unchanged. It also became apparent from the voting and the discussion, that a new spinal lesion would override even clinically significant improvement in all other parameters to lead to a vote of not clinically improved or not MCID. Pain and improvement of sacroiliac joint (SIJ) arthritis or spinal lesion were considered priorities by voters. However, in discussion of certain cases, the combination of CNO lesion count and PHGA or PAGA appeared to take precedence over pain. Thirty percent of improvements in core variables and improvement of CDAS by 3 were considered meaningful by providers and consensus data. This was consistent with other rheumatological diseases as well as data from a prior study by Wu et al (14). Further argument for use of MCID is that this threshold of minimal improvement is simpler to calculate than the Pediatric CNO response (PedCNO30) score which requires at least 30% improvement in at least three of five core set variables (ESR, number of radiologic lesions, PHGA, PAGA and the Childhood Health Assessment Questionnaire (CHAQ)), with no more than one of the remaining variables deteriorating by more than 30%.

Additionally, the main limitation of the PedCNO score in the context of disease activity is that it reports only relative changes in disease activity, even though the score bases are absolute measurements. Moreover, the availability of repeat whole-body MRI (WB-MRI) analyses, as requested, is not guaranteed in all centers (23).

A major strength of this study is the use of a large prospective registry with real-world data for cohort creation for voting. Some missing data were imputed, but most variables were available. Internally inconsistent cases were not included in the final analysis. Members of the expert panel reviewed >1800 cases in total. Cases which did not reach consensus virtually were discussed further during the face-to-face consensus meeting. As part of nominal group technique, members of the expert panel including patients with CNO, caregivers of pediatric patients with CNO, and pediatric rheumatologists interacted during discussions about voting decisions. While these decisions may not be entirely in accordance with their initial personal view, they were aligned with choices that they were willing to accept after discussion. Through the intensive process, panelists recontextualized and opinions evolved. For example, pediatric rheumatologists became more cognizant of pain levels after hearing experiences from patients, while patients weighed the number of MRI lesions less heavily through discussions from the pediatric rheumatologists. Study limitations included some assumptions made for each clinical scenario. For example, for voting purposes the pain level was deemed pain attributable only to CNO. However, clinically, differentiating pain level scores due to CNO or from co-morbid pain amplification syndrome or from mechanical causes of pain is difficult as inflammatory markers are not uniformly elevated in CNO with active inflammatory lesions (8, 24). Whole-body MRI is helpful to differentiate active CNO from amplified musculoskeletal pain syndromes (25). In order to address the possibility of amplified musculoskeletal pain contributing to CDAS scores, we also added a caveat that if CDAS ≥ 3, WB-MRI must show at least one CNO lesion or arthritis to qualify as non-MDA or flare (Figures 3B and 4B, respectively).These additional criteria may help avoid the use of ineffective drugs in patients with pain amplification and allow providers to recommend other modalities such as physical therapy to address pain. We also developed these definitions based solely on the musculoskeletal manifestations of CNO. Other organ manifestations (including skin or gut manifestations) are not included in these scenarios as it is not known whether they parallel MSK manifestations. We also recognized that the concept of MDA, flare, and MCID may or may not necessitate actual treatment change as treatment decisions often include considerations beyond just clinical status (e.g. parental concern for treatment safety, prior drug side effects, insurance issues). Expert panelists stated that the location and severity of lesions on the MRI would normally influence this decision about MDA and treatment when the MRI count is elevated. That information was not provided in the patient profile other than presence of spinal or SIJ lesion. For MCID voting, we also assumed the MRI was completed at the same time as the other parameters, which is not always the case in real-life. We also assumed that the pain scores in patient profiles were the *average* pain over a period of time. Thus, during this initial phase of development of these terms, the definitions should be limited to research settings. When these preliminary definitions have been further validated in different settings, it could potentially be used in clinical practice. Observational registries such as Eurofever (6) and the German National Pediatric Rheumatology Registry (26) can provide real-world data for external validations of these definitions. Additionally, these definitions should be regularly reviewed as treatment options become available for CNO.

## Conclusion

Through a collaborative, data-driven approach with input from patients, caregivers, and pediatric rheumatologists we accomplished our aims to develop definitions for MDA, flare, and MCID in CNO. A Delphi survey with responses from around the world identified important variables to define MDA, flare, and MCID in patients with CNO. Though initial priorities selected by providers differed from those selected by patients/caregivers for MDA/flare, through intensive nominal group technique, consensus was reached for a large proportion of cases. Absence of active MRI lesions, MRI-confirmed sacroiliitis, and active clinical lesions were necessary to meet criteria for MDA. In contrast, presence of these parameters led voters to classify cases as representative of disease flare. Thirty percent of improvements in pain level and improvement of CDAS by 3 were considered meaningful by providers and consensus data for MCID. Thus, we developed preliminary definitions of MDA, flare, and MCID in patients with CNO that represent useful treatment target states and are proposed for inclusion as outcome measures in future observational studies and clinical trials in patients with CNO.

## Data Availability

All data produced in the present study are available upon reasonable request to the authors

## Acknowledgements and Affiliations

The authors wish to acknowledge the CARRA CNO Workgroup for support of this project. The authors also wish to acknowledge CARRA and the ongoing Arthritis Foundation financial support of CARRA. The Arthritis Foundation and CARRA provided bridge funding to YZ. Kaila’s Komfort provided funding for the consensus conference. Findings from this study were presented as abstracts at American College of Rheumatology Convergence 2025 and will be published in an online supplement of the scientific journal, *Arthritis & Rheumatology*.

## Ethics/Institutional Review Board Approval

Institutional Review Board of University of Washington gave approval for this work in IRB #5217 and #1232.

