## Supplement 1 for "Development and Validation of Minimal Disease Activity, Disease Flare, and Minimal Clinically Important Difference for Children with Chronic Nonbacterial Osteomyelitis Using a Consensus and Data-Driven Approach"

### CNO Patient and Caregiver Survey

#### Demographics

| | |
|---|---|
| Please indicate which of the following you are: | ◯ Patient who was diagnosed with CNO as a child (17 and under)<br>◯ Patient who was diagnosed with CNO as an adult (18 and over)<br>◯ Caregiver of a patient diagnosed with CNO as a child<br>◯ Caregiver of a patient diagnosed with CNO as an adult |
| Where do you live? | ◯ North America ______<br>◯ South America<br>◯ Africa<br>◯ Europe<br>◯ Asia<br>◯ Australia/Oceania |
| How many years ago were you diagnosed with CNO? | ◯ less than 5<br>◯ 5-10<br>◯ 11-15<br>◯ 16-20<br>◯ more than 20 |
| How many years have you been caring for someone diagnosed with CNO? | ◯ less than 5<br>◯ 5-10<br>◯ 11-15<br>◯ 16-20<br>◯ more than 20 |
| Have you ever taken medication for CNO? | ◯ Yes<br>◯ No |
| Do you have experience helping the patient take medications to treat CNO? | ◯ Yes<br>◯ No |
| Have you ever had a disease flare? A flare happens during a time when symptoms are under control and a medication is stopped, but then there is an increase or return of symptoms. During a flare the patient feels so unwell that they decide with their doctor to start medications again. | ◯ Yes<br>◯ No |
| Have you helped care for the patient during a disease flare? A flare happens during a time when symptoms are under control and a medication is stopped, but then there is an increase or return of symptoms. During a flare the patient feels so unwell that they decide with their doctor to start medications again. | ◯ Yes<br>◯ No |

#### Minimal Disease Activity (MDA)

Minimal disease activity (MDA) means that a patient feels pretty good or well enough that neither the patient nor their doctor thinks treatment needs to change right now.

What signs or features do you think show MDA for CNO? Please select all that apply. Assume that the features are only related to a patient's CNO disease. Don't choose anything you don't think is important. Please be sure to fill out the text box for every feature you choose. If you have other ideas that aren't listed, feel free to add them at the bottom.

______

**Please rank the MDA (minimal disease activity) signs you picked from the MOST IMPORTANT to the LEAST IMPORTANT.**

**Depending on how many signs you picked, some numbers may be left blank.**

| | Most Important (1) | (2) | (3) | (4) | (5) | (6) | (7) | (8) | (9) | (10) | (11) | (12) | (13) | Least Important (14) |
|---|---|---|---|---|---|---|---|---|---|---|---|---|---|---|
| Patient or caregiver reports pain at [pt_pain_fam] or lower. | ○ | ○ | ○ | ○ | ○ | ○ | ○ | ○ | ○ | ○ | ○ | ○ | ○ | ○ |
| Patient or caregiver reports the level of how much CNO affects the patient's overall state of well-being at [pt_global_fam] or lower. | ○ | ○ | ○ | ○ | ○ | ○ | ○ | ○ | ○ | ○ | ○ | ○ | ○ | ○ |
| Total number of body parts that have pain or swelling caused by CNO is [lesion_ct_fam] or less | ○ | ○ | ○ | ○ | ○ | ○ | ○ | ○ | ○ | ○ | ○ | ○ | ○ | ○ |
| Doctor assesses the level of how much CNO affects the patient's overall state of well-being at [pga_fam] or lower. | ○ | ○ | ○ | ○ | ○ | ○ | ○ | ○ | ○ | ○ | ○ | ○ | ○ | ○ |
| Total number of body parts that have pain or swelling caused by CNO that are also seen on an MRI is [mri_ct_fam] or lower | ○ | ○ | ○ | ○ | ○ | ○ | ○ | ○ | ○ | ○ | ○ | ○ | ○ | ○ |
| Total number of body parts that are noted on an MRI to have CNO that may or may not match up with pain or swelling the patient is feeling is [total_ct_fam] or lower | ○ | ○ | ○ | ○ | ○ | ○ | ○ | ○ | ○ | ○ | ○ | ○ | ○ | ○ |
| Blood tests that show inflammation in the body at levels that are [esr_fam] | ○ | ○ | ○ | ○ | ○ | ○ | ○ | ○ | ○ | ○ | ○ | ○ | ○ | ○ |
| No active arthritis - arthritis is when joints are inflamed | ○ | ○ | ○ | ○ | ○ | ○ | ○ | ○ | ○ | ○ | ○ | ○ | ○ | ○ |
| The patient can move all their body parts without any issues or pain | ○ | ○ | ○ | ○ | ○ | ○ | ○ | ○ | ○ | ○ | ○ | ○ | ○ | ○ |
| No fevers | ○ | ○ | ○ | ○ | ○ | ○ | ○ | ○ | ○ | ○ | ○ | ○ | ○ | ○ |
| [another_item1_fam] | ○ | ○ | ○ | ○ | ○ | ○ | ○ | ○ | ○ | ○ | ○ | ○ | ○ | ○ |
| [another_item2_fam] | ○ | ○ | ○ | ○ | ○ | ○ | ○ | ○ | ○ | ○ | ○ | ○ | ○ | ○ |
| [another_item3_fam] | ○ | ○ | ○ | ○ | ○ | ○ | ○ | ○ | ○ | ○ | ○ | ○ | ○ | ○ |
| [another_item4_fam] | ○ | ○ | ○ | ○ | ○ | ○ | ○ | ○ | ○ | ○ | ○ | ○ | ○ | ○ |

#### Medication Timelines

After a patient feels completely better (which means no pain, no trouble moving, and no growth problems), while taking a medication that is working, how long would you keep using each type of medication before stopping?

Since different families and doctors may feel comfortable for different amounts of time, please pick both the shortest and longest time you would use each treatment. Please enter 0 if you would stop the medication immediately when the patient feels better.

NSAID such as ibuprofen, naproxen, indomethacine, Celebrex, diclofenac ______ ______ months as the shortest time ______ months as the longest time
DMARDsuch as methotrexate, leflumomide, sulfasalazine ______ ______ months as the shortest time ______ months as the longest time
Biologic: TNFi such as etanercept, adalimumab, infliximab, golimumab ______ ______ months as the shortest time ______ months as the longest time
Bisphosphonate: Pamidronate ______ ______ months as the shortest time ______ months as the longest time
Bisphosphonate: Zoledronic acid ______ ______ months as the shortest time ______ months as the longest time

#### Disease Flare

This question is asking about disease flare. During a time when symptoms are under control and medication is stopped, disease flare is an increase or return of symptoms. During a flare the patient feels so unwell that they decide with their doctor to start medications again.

Which signs do you think show a disease flare? Assume that the features are only related to a patient's CNO disease. Please select all that apply. Don't choose anything you don't think is important. Please be sure to fill out the text box for every sign you choose. If you have other ideas that aren't listed, please add them at the bottom.

______

**Please rank the disease flare signs you picked from the MOST IMPORTANT to the LEAST IMPORTANT.**

**Depending on how many signs you picked, some numbers may be left blank.**

| | Most important (1) | (2) | (3) | (4) | (5) | (6) | (7) | (8) | (9) | (10) | (11) | (12) | (13) | (14) | (15) | (16) | Least Important (17) |
|---|---|---|---|---|---|---|---|---|---|---|---|---|---|---|---|---|---|
| Patient or caregiver reports pain at [pt_pain_2_fam] or higher | ○ | ○ | ○ | ○ | ○ | ○ | ○ | ○ | ○ | ○ | ○ | ○ | ○ | ○ | ○ | ○ | ○ |
| Patient or caregiver reports the level of how much CNO affects the patient's overall state of well-being at [pt_global_2_fam] or higher. | ○ | ○ | ○ | ○ | ○ | ○ | ○ | ○ | ○ | ○ | ○ | ○ | ○ | ○ | ○ | ○ | ○ |
| Total number of body parts that have pain or swelling caused by CNO is [lesion_ct_2_fam] or more. | ○ | ○ | ○ | ○ | ○ | ○ | ○ | ○ | ○ | ○ | ○ | ○ | ○ | ○ | ○ | ○ | ○ |
| Doctor assesses the level of how much CNO affects the patient's overall state of well-being at [pga_2_fam] or higher. | ○ | ○ | ○ | ○ | ○ | ○ | ○ | ○ | ○ | ○ | ○ | ○ | ○ | ○ | ○ | ○ | ○ |
| Total number of body parts that have pain or swelling caused by CNO that are also seen on an MRI is [mri_ct_2_fam] or higher | ○ | ○ | ○ | ○ | ○ | ○ | ○ | ○ | ○ | ○ | ○ | ○ | ○ | ○ | ○ | ○ | ○ |
| Total number of body parts that are noted on an MRI to have CNO that may or may not match up with pain or swelling the patient is feeling is [total_ct_2_fam] or higher | ○ | ○ | ○ | ○ | ○ | ○ | ○ | ○ | ○ | ○ | ○ | ○ | ○ | ○ | ○ | ○ | ○ |
| Blood tests show inflammation in the body in levels that are [esr_2_fam] | ○ | ○ | ○ | ○ | ○ | ○ | ○ | ○ | ○ | ○ | ○ | ○ | ○ | ○ | ○ | ○ | ○ |
| Active arthritis - arthritis is when joints are inflamed | ○ | ○ | ○ | ○ | ○ | ○ | ○ | ○ | ○ | ○ | ○ | ○ | ○ | ○ | ○ | ○ | ○ |
| Active enthesitis - enthesitis is when the tendons and ligaments in the joints are inflamed | ○ | ○ | ○ | ○ | ○ | ○ | ○ | ○ | ○ | ○ | ○ | ○ | ○ | ○ | ○ | ○ | ○ |
| The patient can't move their body like they normally can | ○ | ○ | ○ | ○ | ○ | ○ | ○ | ○ | ○ | ○ | ○ | ○ | ○ | ○ | ○ | ○ | ○ |
| Fevers | ○ | ○ | ○ | ○ | ○ | ○ | ○ | ○ | ○ | ○ | ○ | ○ | ○ | ○ | ○ | ○ | ○ |

| | | | | | | | | | | | | | | | | | |
|---|---|---|---|---|---|---|---|---|---|---|---|---|---|---|---|---|---|
| Clinical disease activity score (A combination of the first three items is [cdas_fam] or moreas a sum of pain, patient/parent global assessment and clinical lesion count) greater than or equal to [cdas] | ○ | ○ | ○ | ○ | ○ | ○ | ○ | ○ | ○ | ○ | ○ | ○ | ○ | ○ | ○ | ○ | ○ |
| [another_item5_fam] | ○ | ○ | ○ | ○ | ○ | ○ | ○ | ○ | ○ | ○ | ○ | ○ | ○ | ○ | ○ | ○ | ○ |
| [another_item6_fam] | ○ | ○ | ○ | ○ | ○ | ○ | ○ | ○ | ○ | ○ | ○ | ○ | ○ | ○ | ○ | ○ | ○ |
| [another_item7_fam] | ○ | ○ | ○ | ○ | ○ | ○ | ○ | ○ | ○ | ○ | ○ | ○ | ○ | ○ | ○ | ○ | ○ |
| [another_item8_fam] | ○ | ○ | ○ | ○ | ○ | ○ | ○ | ○ | ○ | ○ | ○ | ○ | ○ | ○ | ○ | ○ | ○ |
