## Supplement 2 for "Development and Validation of Minimal Disease Activity, Disease Flare, and Minimal Clinically Important Difference for Children with Chronic Nonbacterial Osteomyelitis Using a Consensus and Data-Driven Approach"

### CNO Health Care Provider Survey

#### Demographics

Please indicate which of the following you are:

- ◯ Pediatric (or med/peds) rheumatology attending
- ◯ Pediatric (or med/peds) rheumatology fellow
- ◯ Pediatric rheumatology Nurse Practitioner or Physician Assistant
- ◯ Other health care related person

Please explain your role:

__________________________________

How many years have you practiced pediatric rheumatology (including fellowship or other subspecialty training)?

- ◯ < 5
- ◯ 5-10
- ◯ 11-15
- ◯ 16-20
- ◯ >20

Have you ever been involved in the care of children with CNO?

- ◯ Yes
- ◯ No

How many children with CNO do you personally care for in a typical calendar year?

- ◯ < 5
- ◯ 5-10
- ◯ 11-15
- ◯ 16-20
- ◯ >20

How confident are you in treating children with CNO?

- ◯ Completely confident
- ◯ Very confident
- ◯ Moderately confident
- ◯ Somewhat confident
- ◯ Not at all confident

#### Minimal Disease Activity (MDA)

Minimal disease activity (MDA) refers to a state when a patient feels completely or reasonably well where the patient/caregiver and medical provider do not feel the need to change medical therapies at that time.

Which of the following features define(s) MDA of musculoskeletal symptoms to you? Assume that the features are only related to a patient's CNO disease.

SELECT ALL THAT APPLY by checking the checkbox. Do NOT choose those you do not consider necessary. Please be sure to both check the box and input the value within the same response.

If the feature is not listed, please add the features you consider at the end (as many as you like). Consider each item separately and individually.

______

**Please rank your indicated MDA features from the MOST IMPORTANT to the LEAST IMPORTANT.**

**If you did not select every feature in the last question, some numbers will be left blank.**

| | Most Important (1) | (2) | (3) | (4) | (5) | (6) | (7) | (8) | (9) | (10) | (11) | (12) | (13) | Least Important (14) |
|---|---|---|---|---|---|---|---|---|---|---|---|---|---|---|
| Patient/caregiver pain score (0-10) less than or equal to [pt_pain] | ○ | ○ | ○ | ○ | ○ | ○ | ○ | ○ | ○ | ○ | ○ | ○ | ○ | ○ |
| Patient/caregiver global assessment of rheumatic (CNO) disease (0-10) less than or equal to [pt_global] | ○ | ○ | ○ | ○ | ○ | ○ | ○ | ○ | ○ | ○ | ○ | ○ | ○ | ○ |
| A total CNO clinical lesion count of less than or equal to [lesion_ct] | ○ | ○ | ○ | ○ | ○ | ○ | ○ | ○ | ○ | ○ | ○ | ○ | ○ | ○ |
| Physician global assessment (0-10) less than or equal to [pga] | ○ | ○ | ○ | ○ | ○ | ○ | ○ | ○ | ○ | ○ | ○ | ○ | ○ | ○ |
| Symptomatic MRI lesion count(s) less than or equal to [mri_ct] | ○ | ○ | ○ | ○ | ○ | ○ | ○ | ○ | ○ | ○ | ○ | ○ | ○ | ○ |
| Total MRI lesion count(s) including asymptomatic lesions less than or equal to [total_ct] | ○ | ○ | ○ | ○ | ○ | ○ | ○ | ○ | ○ | ○ | ○ | ○ | ○ | ○ |
| Inflammatory markers, erythrocyte sedimentation rate (ESR), normal or less than [esr] | ○ | ○ | ○ | ○ | ○ | ○ | ○ | ○ | ○ | ○ | ○ | ○ | ○ | ○ |
| No active arthritis | ○ | ○ | ○ | ○ | ○ | ○ | ○ | ○ | ○ | ○ | ○ | ○ | ○ | ○ |
| No functional limitation of joints/limbs | ○ | ○ | ○ | ○ | ○ | ○ | ○ | ○ | ○ | ○ | ○ | ○ | ○ | ○ |
| No fevers | ○ | ○ | ○ | ○ | ○ | ○ | ○ | ○ | ○ | ○ | ○ | ○ | ○ | ○ |
| [another_item1] | ○ | ○ | ○ | ○ | ○ | ○ | ○ | ○ | ○ | ○ | ○ | ○ | ○ | ○ |
| [another_item2] | ○ | ○ | ○ | ○ | ○ | ○ | ○ | ○ | ○ | ○ | ○ | ○ | ○ | ○ |
| [another_item3] | ○ | ○ | ○ | ○ | ○ | ○ | ○ | ○ | ○ | ○ | ○ | ○ | ○ | ○ |
| [another_item4] | ○ | ○ | ○ | ○ | ○ | ○ | ○ | ○ | ○ | ○ | ○ | ○ | ○ | ○ |

#### Medication Timelines

After a patient achieves remission (defined as the absence of pain, functional limitations, or growth disturbances), how long would you continue each individual effective treatment (assuming each treatment alone is sufficient) before stopping?

Because there is a range of the duration each family and/or physician would be comfortable with, we ask you to choose both the shortest duration and the longest duration for each treatment. Please enter 0 if you would stop the medication immediately upon remission.

NSAID such as ibuprofen, naproxen, indomethacine, Celebrex, diclofenac ______ ______ months as the shortest duration ______ months as the longest duration
DMARDsuch as methotrexate, leflumomide, sulfasalazine ______ ______ months as the shortest duration ______ months as the longest duration
Biologic: TNFi such as etanercept, adalimumab, infliximab, golimumab ______ ______ months as the shortest duration ______ months as the longest duration
Bisphosphonate: Pamidronate ______ ______ months as the shortest duration ______ months as the longest duration
Bisphosphonate: Zoledronic acid ______ ______ months as the shortest duration ______ months as the longest duration

#### Disease Flare

When a medication is stopped during medicated remission, which of the following features define(s) a disease flare or recurrence of musculoskeletal symptoms to you? This refers to a state when a patient feels so unwell that a patient and medical provider need to restart medications.

SELECT ALL THAT APPLY, do NOT choose those you do not consider necessary. Assume that the features are only related to a patient's CNO disease.

Be sure to complete the text box for all fetures selected. If the feature is not listed, please add the features you consider (as many as you like). Consider each item separately and individually.

______

**Please rank your indicated disease flare features from the MOST IMPORTANT to the LEAST IMPORTANT.**

**If not all features were selected in the previous question, some numbers will be left blank**

| | Most important (1) | (2) | (3) | (4) | (5) | (6) | (7) | (8) | (9) | (10) | (11) | (12) | (13) | (14) | (15) | (16) | Least Important (17) |
|---|---|---|---|---|---|---|---|---|---|---|---|---|---|---|---|---|---|
| Patient/caregiver pain score (0-10) greater than or equal to [pt_pain_2] | ○ | ○ | ○ | ○ | ○ | ○ | ○ | ○ | ○ | ○ | ○ | ○ | ○ | ○ | ○ | ○ | ○ |
| Patient/caregiver global assessment of rheumatic (CNO) disease (0-10) greater than or equal to [pt_global_2] | ○ | ○ | ○ | ○ | ○ | ○ | ○ | ○ | ○ | ○ | ○ | ○ | ○ | ○ | ○ | ○ | ○ |
| Total number of CNO clinical lesion count greater than or equal to [lesion_ct_2] | ○ | ○ | ○ | ○ | ○ | ○ | ○ | ○ | ○ | ○ | ○ | ○ | ○ | ○ | ○ | ○ | ○ |
| Physician global assessment (0-10) greater than or equal to [pga_2] | ○ | ○ | ○ | ○ | ○ | ○ | ○ | ○ | ○ | ○ | ○ | ○ | ○ | ○ | ○ | ○ | ○ |
| Symptomatic MRI lesion count(s) greater than or equal to [mri_ct_2] | ○ | ○ | ○ | ○ | ○ | ○ | ○ | ○ | ○ | ○ | ○ | ○ | ○ | ○ | ○ | ○ | ○ |
| Total MRI lesions count(s) including asymptomatic lesions greater than or equal to [total_ct_2] | ○ | ○ | ○ | ○ | ○ | ○ | ○ | ○ | ○ | ○ | ○ | ○ | ○ | ○ | ○ | ○ | ○ |
| Inflammation marker, erythrocyte sedimentation rate (ESR), contributed by CNO, abnormal or greater than contributed by CNO [esr_2] | ○ | ○ | ○ | ○ | ○ | ○ | ○ | ○ | ○ | ○ | ○ | ○ | ○ | ○ | ○ | ○ | ○ |
| Active arthritis | ○ | ○ | ○ | ○ | ○ | ○ | ○ | ○ | ○ | ○ | ○ | ○ | ○ | ○ | ○ | ○ | ○ |
| Active enthesitis | ○ | ○ | ○ | ○ | ○ | ○ | ○ | ○ | ○ | ○ | ○ | ○ | ○ | ○ | ○ | ○ | ○ |
| Functional limitation of joints/limbs | ○ | ○ | ○ | ○ | ○ | ○ | ○ | ○ | ○ | ○ | ○ | ○ | ○ | ○ | ○ | ○ | ○ |
| Fevers | ○ | ○ | ○ | ○ | ○ | ○ | ○ | ○ | ○ | ○ | ○ | ○ | ○ | ○ | ○ | ○ | ○ |
| Clinical disease activity score (CDAS as a sum of pain, patient/parent global assessment and clinical lesion count) greater than or equal to [cdas] | ○ | ○ | ○ | ○ | ○ | ○ | ○ | ○ | ○ | ○ | ○ | ○ | ○ | ○ | ○ | ○ | ○ |
| [another_item5] | ○ | ○ | ○ | ○ | ○ | ○ | ○ | ○ | ○ | ○ | ○ | ○ | ○ | ○ | ○ | ○ | ○ |
| [another_item6] | ○ | ○ | ○ | ○ | ○ | ○ | ○ | ○ | ○ | ○ | ○ | ○ | ○ | ○ | ○ | ○ | ○ |

| | | | | | | | | | | | | | | | | | |
|---|---|---|---|---|---|---|---|---|---|---|---|---|---|---|---|---|---|
| [another_item7] | ○ | ○ | ○ | ○ | ○ | ○ | ○ | ○ | ○ | ○ | ○ | ○ | ○ | ○ | ○ | ○ | ○ |
| [another_item8] | ○ | ○ | ○ | ○ | ○ | ○ | ○ | ○ | ○ | ○ | ○ | ○ | ○ | ○ | ○ | ○ | ○ |

#### Minimal Clinical Important Difference (MCID)

In order to design a clinical trial we need to determine an estimation of meaningful improvement of a disease caused by an effective medication. This is called the minimal clinical important difference (MCID). MCID is a way of measuring if a certain medication is working. It can be a set of criteria or a single measurement. Please note that a benchmark of a certain number may not be reachable if the initial disease is mild so please express such change in both absolute (specific numbers) and relative (%) values. Which of the following should be considered for defining MCID?

SELECT ALL THAT APPLY, do NOT choose those you do not consider necessary. If the feature is not listed, please add the features you consider (as many as you like). Consider each item separately and individually.

__________

**Please rank your indicated MCID features from the MOST IMPORTANT to the LEAST IMPORTANT.**

| | Most Important (1) | (2) | (3) | (4) | (5) | (6) | (7) | (8) | (9) | (10) | (11) | (12) | Least Important (13) |
|---|---|---|---|---|---|---|---|---|---|---|---|---|---|
| Patient/caregiver pain score (0-10) improved by greater than or equal to [pt_pain_abs] or [pt_pain_per]% | ○ | ○ | ○ | ○ | ○ | ○ | ○ | ○ | ○ | ○ | ○ | ○ | ○ |
| Patient/caregiver global assessment of rheumatic (CNO) disease (0-10) improved by than or equal to [pt_global_abs] or [pt_global_per]% | ○ | ○ | ○ | ○ | ○ | ○ | ○ | ○ | ○ | ○ | ○ | ○ | ○ |
| A total CNO clinical lesion count improved by greater than or equal to [lesion_ct_abs] or [lesion_ct_per]% | ○ | ○ | ○ | ○ | ○ | ○ | ○ | ○ | ○ | ○ | ○ | ○ | ○ |
| Physician global assessment (0-10) improved by greater than or equal to [pga_abs] or [pga_per]% | ○ | ○ | ○ | ○ | ○ | ○ | ○ | ○ | ○ | ○ | ○ | ○ | ○ |
| Symptomatic MRI lesion count(s) improved by greater than or equal to [mri_ct_abs] or [mri_ct_per]% | ○ | ○ | ○ | ○ | ○ | ○ | ○ | ○ | ○ | ○ | ○ | ○ | ○ |
| Total MRI lesion count(s) including asymptomatic lesions improved by greater than or equal to [total_ct_abs] or [total_ct_per]% | ○ | ○ | ○ | ○ | ○ | ○ | ○ | ○ | ○ | ○ | ○ | ○ | ○ |
| Inflammatory markers, erythrocyte sedimentation rate (ESR), contributed by CNO improved by greater than [esr_abs] or [esr_per]% | ○ | ○ | ○ | ○ | ○ | ○ | ○ | ○ | ○ | ○ | ○ | ○ | ○ |
| Active arthritis with joint count improved by greater than or equal to [arth_abs] or [arth_per]% | ○ | ○ | ○ | ○ | ○ | ○ | ○ | ○ | ○ | ○ | ○ | ○ | ○ |
| Clinical disease activity score improved by greater than or equal to [cdas_abs] or [cdas_per]% | ○ | ○ | ○ | ○ | ○ | ○ | ○ | ○ | ○ | ○ | ○ | ○ | ○ |
| [another_item9] | ○ | ○ | ○ | ○ | ○ | ○ | ○ | ○ | ○ | ○ | ○ | ○ | ○ |
| [another_item10] | ○ | ○ | ○ | ○ | ○ | ○ | ○ | ○ | ○ | ○ | ○ | ○ | ○ |

| | | | | | | | | | | | | | |
|---|---|---|---|---|---|---|---|---|---|---|---|---|---|
| [another_item11] | ○ | ○ | ○ | ○ | ○ | ○ | ○ | ○ | ○ | ○ | ○ | ○ | ○ |
| [another_item12] | ○ | ○ | ○ | ○ | ○ | ○ | ○ | ○ | ○ | ○ | ○ | ○ | ○ |
